# Lung water density is increased in patients at risk of heart failure and is largely independent of conventional CMR measures

**DOI:** 10.1101/2024.05.31.24308269

**Authors:** Nithin R. Iyer, Jennifer A. Bryant, Thu-Thao Le, Justin G. Grenier, Richard B. Thompson, Calvin W-L. Chin, Martin Ugander

## Abstract

**Aims:** Non-invasive methods to quantify pulmonary congestion are lacking in clinical practice. Cardiovascular magnetic resonance (CMR) lung water density (LWD) mapping is accurate, reproducible, and has prognostic value. However, it is not known whether LWD is associated with routinely acquired CMR parameters.

**Methods and Results:** This was a prospective, observational cohort including healthy controls and patients at risk of heart failure. Lung water density was measured using CMR with a free-breathing short echo time 3D cartesian gradient-echo sequence with a respiratory navigator at 1.5T. Associations were assessed between LWD, lung water volume (LWV) and cardiac volumes, left ventricular (LV) mass and function, myocardial native T1 and extracellular volume fraction (ECV). In patients at risk for heart failure (n=155), LWD was greater than in healthy controls (n=15) (30.4±5.0 vs 27.2±4.3%, p=0.02). Using receiver operating characteristic analysis, the optimal cut-off for LWD was 27.6% to detect at-risk patients (sensitivity 72%, specificity 73%, positive likelihood ratio 2.7, inverse negative likelihood ratio 2.6). LWD was univariably associated with body mass index (BMI), hypertension, right atrial (RA) area and LV mass (p<0.05 for all). In multivariable linear regression, only BMI remained associated with LWD (R^2^=0.32, p<0.001). In a separate multivariable analysis including only CMR markers, only RA area remained associated with LWD (R^2^=0.05, p=0.006).

**Conclusion:** LWD is increased in patients at risk for heart failure compared to controls, and is only weakly explained by conventional CMR measures. LWD provides diagnostic information which is largely independent of conventional CMR measures.

## Background

Pulmonary oedema is the accumulation of extravascular fluid in the lung parenchyma. Cardiogenic pulmonary oedema occurs in heart failure due to an increase in hydrostatic pressure in the pulmonary capillaries resulting from increased left atrial pressure and is associated with a poor long-term prognosis [1–3]. Current methods to assess pulmonary oedema are either invasive (transpulmonary thermodilution), semi-quantitative (chest x-Ray and lung ultrasound) or require the use of ionising radiation (nuclear imaging and computed tomography).

Cardiovascular magnetic resonance imaging (CMR) is a non-invasive imaging modality that is well-suited to quantify lung water content without the need for ionising radiation, and has shown good correlation with gravimetric lung weight in animal models [4]. In human studies, estimated lung water density (LWD) in a heart failure cohort measured with the widely available single-shot fast spin echo (HASTE) pulse sequence was shown to be correlated with invasively measured left-sided filling pressures and predictive of cardiovascular outcomes [5]. More recently, custom non-Cartesian gradient-echo pulse sequences with short echo times have been shown to provide three-dimensional LWD acquisitions in a single breath-hold or respiratory navigator free-breathing acquisitions [2, 6–8]. These methods have addressed practical challenges that must be overcome for accurate estimation of LWD including the low signal-to-noise ratio due to low proton density in the lungs, rapid T2* signal decay, large required field of view and three-dimensional (3D) spatial coverage, respiratory motion, the need for a reliable signal reference and MRI signal normalization [2]. However, these custom methods are not widely available.

The goals of the current study, therefore, were to: 1) assess feasibility of LWD measurement in healthy controls and patients at risk of heart failure using a widely available 3D Cartesian gradient echo approach and 2) evaluate the association between LWD and routinely acquired CMR markers including cardiac volumes, mass, function and myocardial extracellular volume fraction.

## Methods

### Study Population

This prospective, observational study cohort consisted of: 1) subjects serially recruited at the time of clinical CMR at the National Heart Centre Singapore from May 2021 through March 2022 and 2) patients with hypertension undergoing CMR as part of ongoing clinical trials at the centre. Asymptomatic controls without known cardiac disease were recruited. Patients were included if they were aged ≥21 years, able to receive intravenous gadolinium-based contrast agents and had no contraindications to CMR. Exclusion criteria included a documented diagnosis of heart failure, missing relevant demographic data, or uninterpretable imaging. Patients were classified as at risk of heart failure if they had a history of coronary artery disease, diabetes mellitus or hypertension, or clinical history or CMR evidence of cardiomyopathy (ischemic, hypertrophic, dilated, amyloidosis). Ethics approval was obtained from the SingHealth Centralized Institutional Review Board and all participants provided written informed consent. The study was conducted in accordance with the principles of the Declaration of Helsinki.

### Cardiovascular magnetic resonance image acquisition

All patients underwent standardized CMR (Siemens Aera 1.5T; Siemens Healthineers, Erlangen, Germany). Balanced steady-state free precession cine images were acquired in the standard long-axis views and a short-axis stack from base to apex, as described previously [9]. LGE images were acquired at 8 min after 0.1 mmol/kg of gadobutrol (Gadovist®, Bayer Pharma AG, Germany) with a phase sensitive inversion-recovery fast gradient echo imaging sequence. The inversion time for optimal myocardial nulling was selected from an inversion time scout sequence. T1 maps were acquired at the basal and mid-ventricular short-axis levels, pre– and 15-min post-contrast with modified Look-Locker inversion-recovery (MOLLI) 5s(3s)3s and 4s(1s)3s(1s)2s acquisition schemes, respectively. Lung water density was measured with a free-breathing short-echo 3D Cartesian gradient-echo sequence with a respiratory navigator. Typical imaging parameters included a matrix size of 96 x 132 x 60 with a 360 mm x 500 mm x 300 mm field of view in the head to foot, right to left and chest to back directions, respectively, 2000Hz/pixel readout bandwidth, echo time (TE) = 0.43 ms, repetition time (TR) = 1.5 ms and flip angle 1°. All images were interpolated to 2.5 mm isotropic resolution. Complete acquisitions were repeated 7 times over 84 seconds of tidal respiration with the insertion of a center of k-space navigator acquisition every 72 lines of k-space (every 108 ms).

### CMR analysis

Image analysis was performed at the National Heart Research Institute Singapore (NHRIS) CMR Core Laboratory using CVI42 software (Circle Cardiovascular Imaging, Calgary, Canada) by trained imaging fellows who were blinded to the clinical data. Cardiac volumes, mass and ejection fraction were analyzed according to standardized protocols [10, 11]. Left ventricular (LV) volumes and mass data were indexed to body surface area. The presence of LGE was assessed qualitatively by two readers according to the recommendations by the Society of CMR [12]. Myocardial native T1 and extracellular volume (ECV) fraction were measured while excluding regions of focal late gadolinium enhancement (LGE), as described previously [9]. Myocardial strain was analysed in the cine images using the Tissue Tracking Plugin [11].

### Lung water analysis

The center of k-space respiratory navigator was used to reconstruct a single 3D lung dataset at the functional residual capacity respiratory phase (minimum lung volume) [2]. Lung water images were processed offline using a custom program in MATLAB (The MathWorks, Natic, MA, USA) [2]. A spatial normalization scheme was applied to eliminate surface coil shading, as previously described [2]. Briefly, all solid tissues surrounding the lungs were fit with a low spatial frequency normalization map that was interpolated over the full field. 3D images were then corrected by division with the normalization map to yield lung voxel intensity in units of relative lung water density (LWD). The lung parenchyma, excluding large vessels, was identified with a region-growing algorithm [2]. Lung volume was calculated as total number of pixels in the lung mask multiplied by the pixel volume. Average LWD was calculated as the mean LWD value from all lung parenchyma pixels. Water volumes in each pixel were calculated as the LWD value multiplied by the voxel volume and the total lung water volume (LWV) as the sum of these volumes over all lung pixels.

### Statistics

Normality was assessed for continuous variables using the Shapiro-Wilk test. Normally distributed data are presented as mean ± standard deviation. Non-normally distributed data are presented as median [interquartile range]. Comparisons were performed for continuous variables using the parametric Student *t*-test or the non-parametric Mann-Whitney U test. Categorical variables are presented as number (percentage) and compared using the χ^2^ test.

Pearson’s rank order test was performed to detect significant correlations between LWD, LWV and routine CMR markers including cardiac volumes, LV mass and ejection fraction, myocardial native T1 and ECV. Multiple linear regression was performed to assess the relationship between LWD, LWV and routinely acquired CMR markers, adjusting for relevant clinical confounders including age, sex, body surface area (BSA), body mass index (BMI), hypertension, diabetes, coronary artery disease, New York Heart Association class and systolic BP. Covariates with a p-value <0.05 in univariable analyses were entered into the multivariable linear regression model to identify independent predictor variables, using forward stepwise selection (probability to remove threshold ≥0.1). A separate multivariable linear regression analysis was performed including only CMR markers. Receiver operating characteristic analysis with Youden’s index was used on the entire cohort to identify an optimal LWD cut-off for discriminating between the control and at risk groups. Diagnostic performance was described using sensitivity, specificity, positive and negative predictive values, and the disease prevalence-independent positive and inverse negative likelihood ratios. Statistical analyses were performed using SPSS Version 28 (Statistical Package for the Social Sciences, International Business Machines, Inc., Armonk, New York, USA) and GraphPad Prism 9.4.1 (GraphPad Software, Inc., San Diego, California, USA). A two-sided p-value <0.05 was considered as statistically significant.

## Results

A flowchart of patient selection is shown in Figure 1. Out of the 50 patients undergoing clinically indicated CMR recruited for the study, 22 were excluded due to an absence of risk factors for heart failure or due to a pre-existing diagnosis of heart failure. In conjunction with 139 patients with at least one risk factor for heart failure enrolled in ongoing clinical trials at our centre, 167 patients underwent CMR including LWD. From this group, 12 patients had missing clinical data or uninterpretable LWD data and were excluded. Out of the 17 healthy volunteers who underwent CMR with LWD, 2 were excluded due to uninterpretable LWD data. In total, 170 subjects (healthy controls, n=15; at risk of heart failure, n= 155) were included in the study cohort.

**Figure 1.**
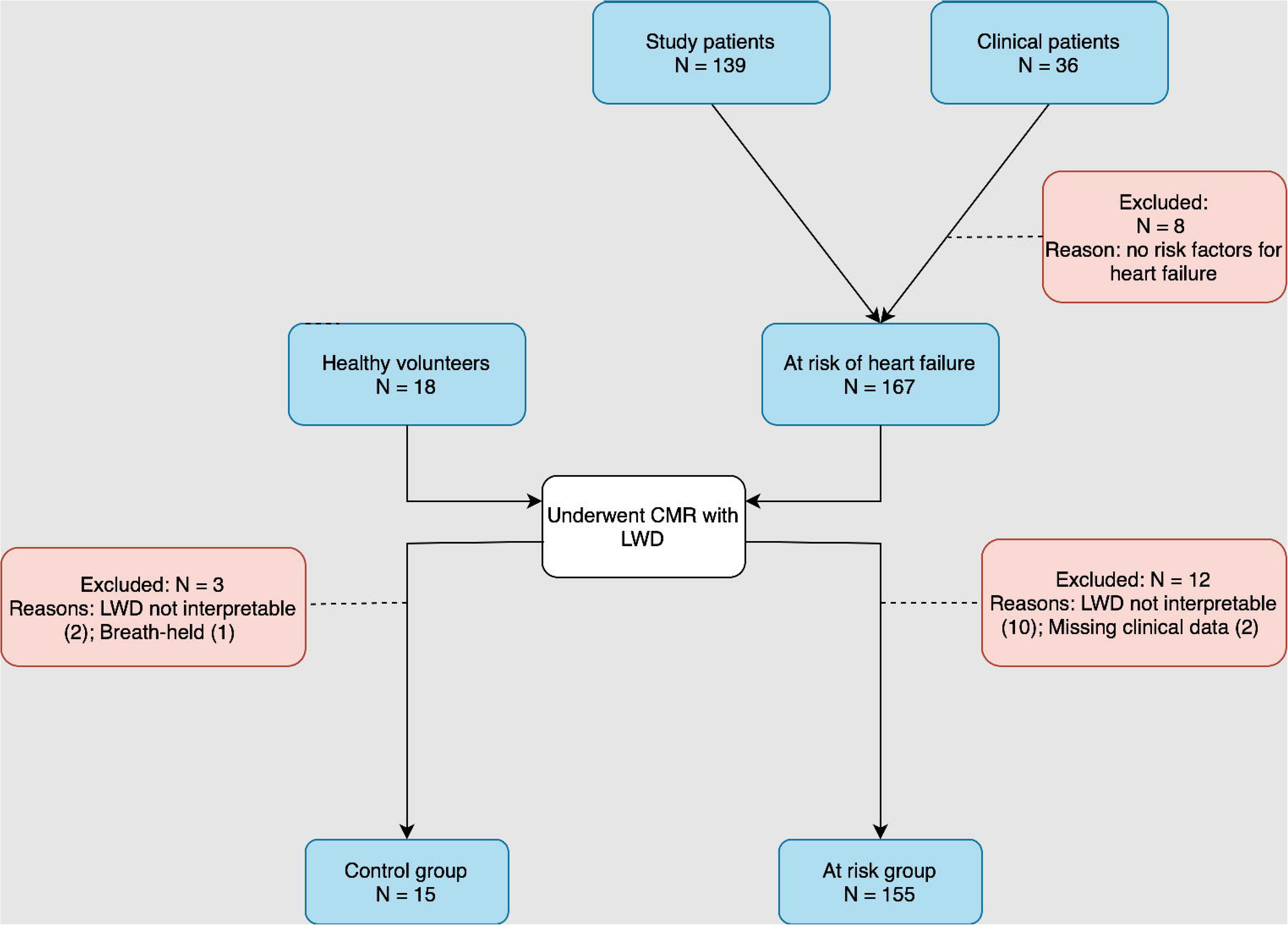
Flow chart of patient inclusion. Abbreviations: CMR: Cardiovascular magnetic resonance; LWD: Lung water density.

## Baseline characteristics

The baseline clinical characteristics are shown in Table 1. Compared to healthy controls, patients at risk of heart failure were older, had higher systolic BP and had higher BMI. At CMR, the at risk group had higher right atrial (RA) area, left atrial (LA) volume and LV mass, worse GLS and higher LWD (Figure 2). LWV did not differ between the two groups.

**Figure 2.**
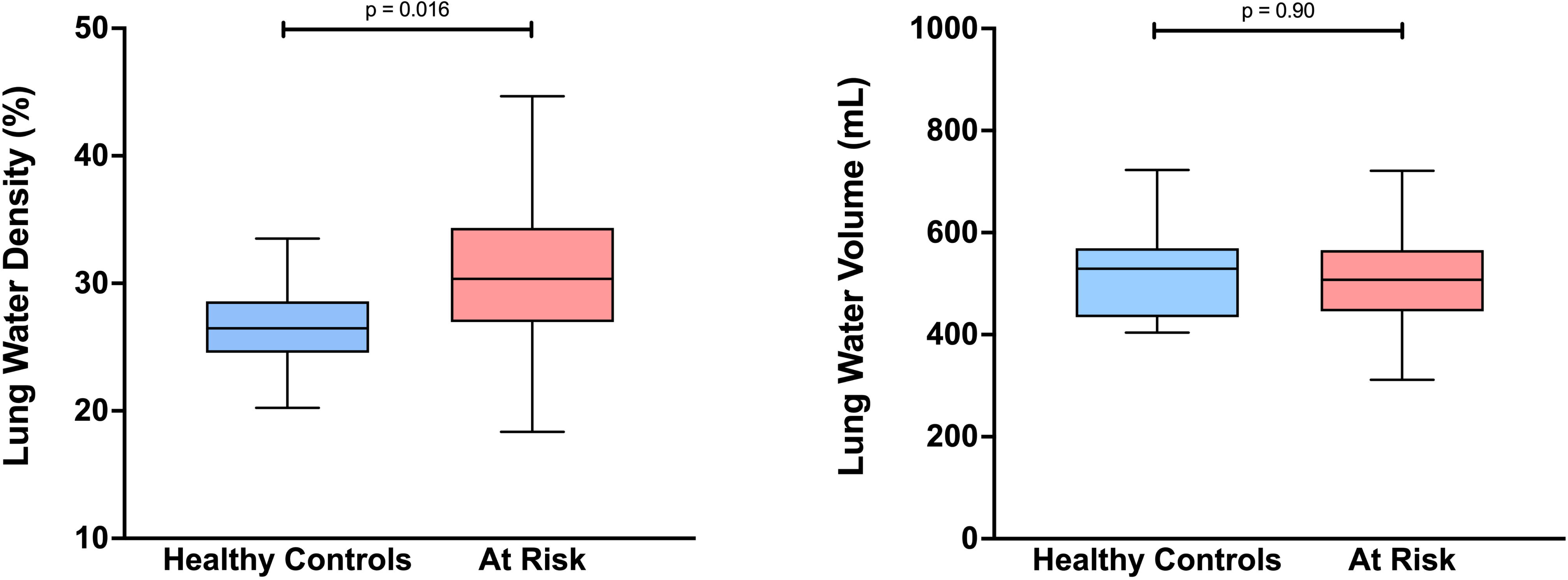
Lung water density and volume according to clinical group: Lung water density was higher in patients at risk of heart failure compared to healthy controls (Panel A); Lung water volume did not differ between healthy controls and patients at risk of heart failure (Panel B). Whiskers were plotted using the Tukey method. Outliers are not shown for clarity.

**Table 1.** Baseline clinical and CMR characteristics of the cohort.

|  | Healthy Controls<br>(n=15) | At risk of heart<br>failure<br>(n=155) | p Value |
| --- | --- | --- | --- |
| **Clinical** |  |  |  |
| Age, years | 48 [38-55] | 60 [50-65] | <0.001 |
| Male sex, n (%) | 8 (53) | 110 (71) | 0.16 |
| Systolic BP, mmHg | 131 ± 19 | 142 ± 18 | 0.006 |
| BSA, m<sup>2</sup> | 1.73 ± 0.25 | 1.86 ± 0.23 | 0.006 |
| BMI, kg/m<sup>2</sup> | 21.6 [19.5-26.5] | 27.0 [23.8-30.1] | <0.001 |
| Co-morbidities, n (%) |  |  |  |
| Hypertension | 0 (0) | 143 (92) |  |
| Diabetes | 0 (0) | 62 (40) |  |
| Coronary artery disease | 0 (0) | 37 (24) |  |
| NYHA Functional Class, n (%) |  |  |  |
| Class I | 15 (100) | 117 (89) |  |
| Class II | 0 (0) | 15 (11) |  |
| **CMR Markers** |  |  |  |
| RA area, m<sup>2</sup> | 12 [11-13] | 20 [16-23] | <0.001 |
| LA volume, mL | 70 [64-81] | 86 [73-104] | 0.013 |
| LV EDV, mL | 131 [120-144] | 134 [112-158] | 0.80 |
| RV EDV, mL | 142 [122-153] | 134 [110-156] | 0.28 |
| LV mass, g | 76 [69-98] | 97 [80-117] | 0.006 |
| LV ejection fraction, % | 57 [56-61] | 58 [54-63] | 0.84 |
| GLS, % | -18.2 ± 1.7 | -15.6 ± 3.3 | 0.004 |
| LGE Type |  |  |  |
| Nil, n (%) | 9 (100) | 88 (57) |  |
| Non-ischaemic, n (%) | 0 (0) | 36 (23) |  |
| Ischaemic, n (%) | 0 (0) | 29 (19) |  |
| Both, n (%) | 0 (0) | 2 (1) |  |
| Native T1, ms | - | 1022 [1008-1038] |  |
| ECV, % | - | 25.6 [23.8-27.2] |  |
| Lung Volume, ml | 1861 [1604-2405] | 1708 [1416-2037] | 0.08 |
| LWD, % | 27.2 ± 4.3 | 30.4 ± 5.0 | 0.02 |
| LWV, mL | 521 ± 94 | 518 ± 98 | 0.90 |

Values are given as median [interquartile range], mean ± SD or number (percentage). **Abbreviations:** BSA: Body surface area; NYHA: New York Heart Association; RA: Right atrial; LA: Left atrial; LV: Left ventricular; EDV: End-diastolic volume; RV: Right ventricular; GLS: Global longitudinal strain; LGE: Late gadolinium enhancement; ECV: Extracellular volume; LWD: Lung water density; LWV: Lung water volume.

## Receiver operating characteristic analysis

Using receiver operating characteristic (ROC) analysis, the area under the curve (AUC) was 0.70 (95% CI 0.58-0.83) for LWD to differentiate between healthy controls and patients at risk of heart failure (Figure 3). The optimal cut-off point for LWD was 27.6% to detect at risk patients with sensitivity 72%, specificity 73%, positive predictive value 96%, negative predictive value 20%, positive likelihood ratio 2.7, and inverse negative likelihood ratio 2.6. For LWV, the AUC was 0.49 (95% CI 0.34-0.65), indicating no ability to discriminate between healthy controls and patients at risk of heart failure.

**Figure 3.**
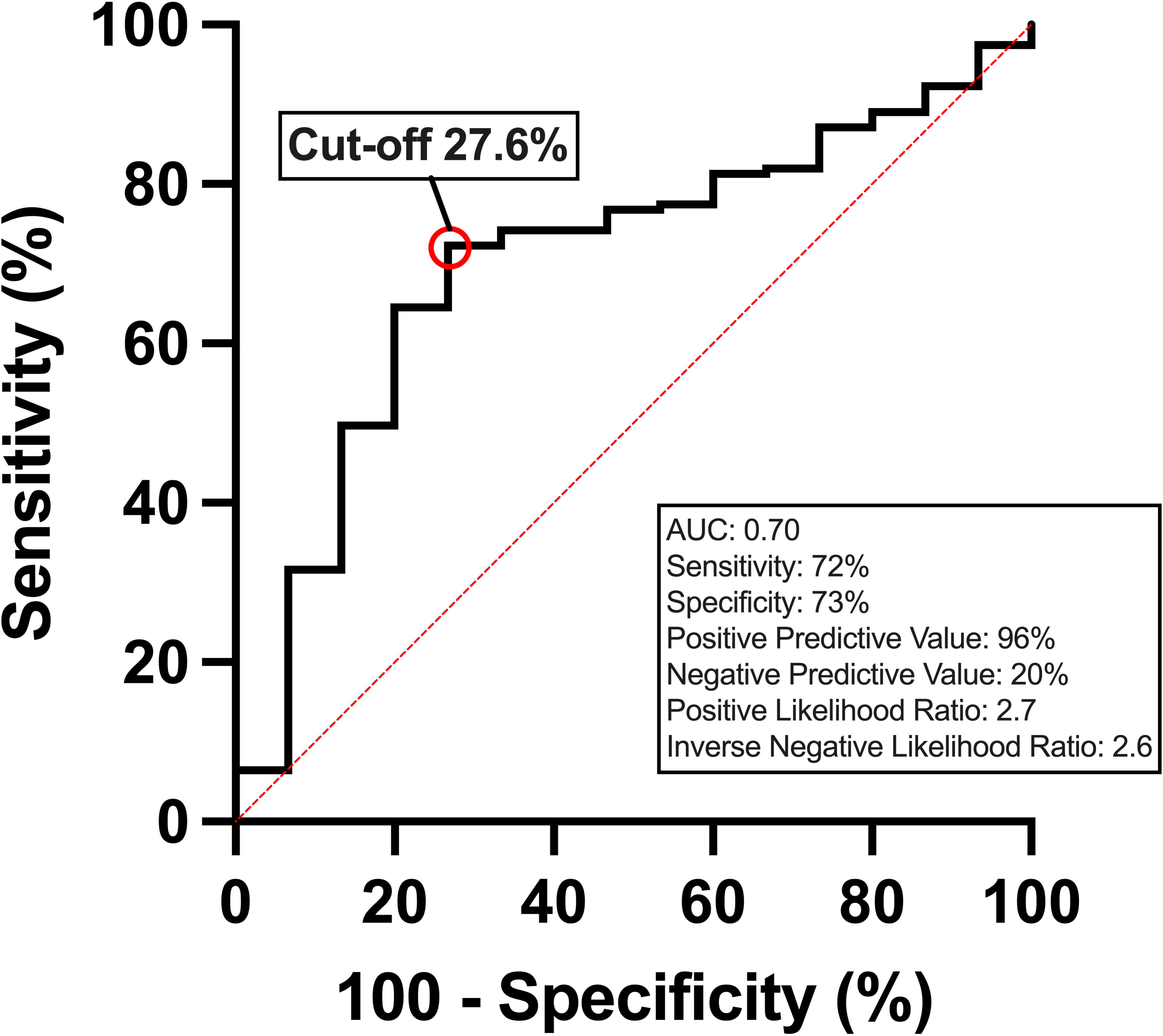
Receiver operating characteristic (ROC) curve and corresponding area under the curve (AUC) describing the diagnostic performance of lung water density (LWD) to detect patients at risk of heart failure.

## Variables associated with LWD

Univariable and multivariable analyses for LWD are shown in Table 2. In univariable linear regression analysis, LWD was associated with BSA, BMI, hypertension, RA area and LV mass (Figure 4). In a stepwise multivariable linear regression analysis including clinical and CMR variables associated with LWD, only BMI remained associated with LWD (model R^2^=0.32, p<0.001). In a separate multivariable linear regression model including only CMR markers associated with LWD, RA area remained associated with LWD (model R^2^=0.05, p=0.006) (Table 3). Representative images showing LWD outputs are shown in Figure 5.

**Figure 4.**
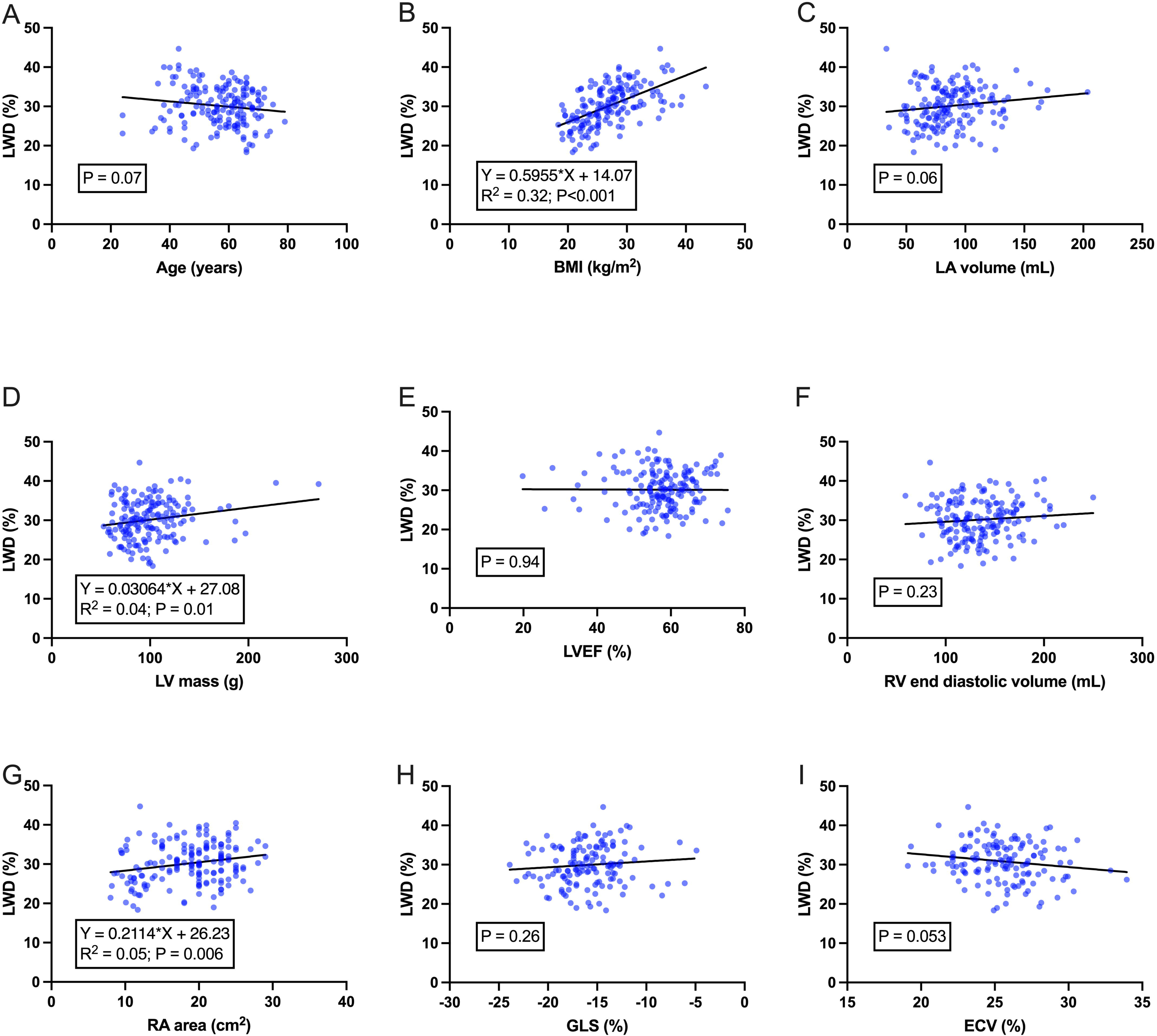
Scatter plots showing univariable relationships between lung water density and clinical or routinely acquired CMR markers. Lung water density was significantly associated with BMI (Panel B), LV mass (Panel D) and RA area (Panel G). The line of best fit is shown for each analysis. Abbreviations: LWD: Lung water density; BMI: Body mass index; LA: Left atrial; LV: Left ventricular; LVEF: Left ventricular ejection fraction; RV: Right ventricular; RA: Right atrial; GLS: Global longitudinal strain; ECV: Extracellular volume fraction.

**Figure 5.**
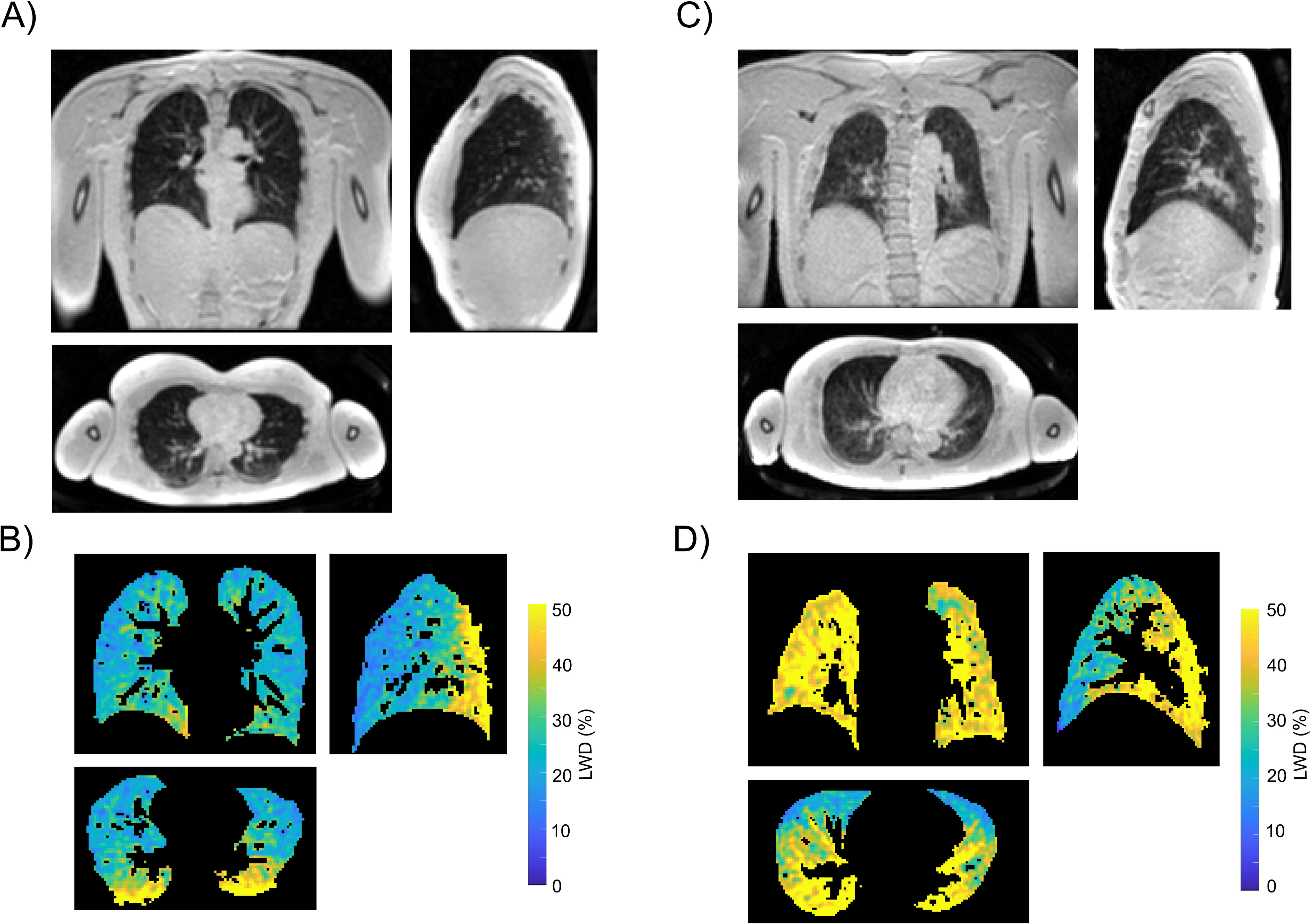
Illustrative lung images. Coronal, sagittal and transverse slices from a three-dimensional Cartesian lung water image in: (A) control subject and (C) patient at risk of heart failure. Lung water density (%) in the lung parenchyma following image normalization and masking for the same slice locations shown in A (Panel B) and C (Panel D) showing globally increased lung water density in the patient.

**Table 2.** Univariable and multivariable linear regression models of clinical and CMR variables associated with LWD. Stepwise selection identified variables associated with LWD with p value <0.05 to enter and remain in the model.

|  | Univariable Model |  | Multivariable Model, stepwise selection (global R<sup>2</sup> = 0.32) |  |
| --- | --- | --- | --- | --- |
|  | R<sup>2</sup> | p Value | t | p Value |
| **Clinical** |  |  |  |  |
| Age, years | 0.019 | 0.07 |  |  |
| Male sex | 0.003 | 0.52 |  |  |
| Body mass index, kg/m<sup>2</sup> | 0.323 | <0.001 | 8.9 | <0.001 |
| Body surface area, m<sup>2</sup> | 0.121 | <0.001 | * |  |
| Hypertension | 0.051 | 0.003 | § |  |
| Diabetes | 0.001 | 0.63 |  |  |
| Coronary artery disease | 0.000 | 0.82 |  |  |
| NYHA Functional Class | 0.001 | 0.69 |  |  |
| Systolic blood pressure, mmHg | 0.018 | 0.09 |  |  |
| **CMR Markers** |  |  |  |  |
| RA area, cm<sup>2</sup> | 0.045 | 0.006 | § |  |
| LA volume, mL | 0.022 | 0.057 |  |  |
| LV EDV, mL | 0.018 | 0.09 |  |  |
| RV EDV, mL | 0.009 | 0.23 |  |  |
| LV mass, g | 0.035 | 0.01 | § |  |
| LV ejection fraction, % | 0.000 | 0.94 |  |  |
| GLS, % | 0.009 | 0.26 |  |  |
| Presence of LGE | 0.025 | 0.054 |  |  |
| Native T1, ms | 0.002 | 0.63 |  |  |
| ECV, % | 0.027 | 0.051 |  |  |
| LWV, mL | 0.001 | 0.74 |  |  |

**Abbreviations:**; NYHA: New York Heart Association; RA: Right atrial; LA: Left atrial; LV: Left ventricular; EDV: End-diastolic volume; RV: Right ventricular; GLS: Global longitudinal strain; LGE: Late gadolinium enhancement; ECV: Extracellular volume; LWV: Lung water volume.
§ Assessed for inclusion in multivariable model but not retained.
\* Excluded from multivariable analysis due to collinearity with BMI.

**Table 3.**
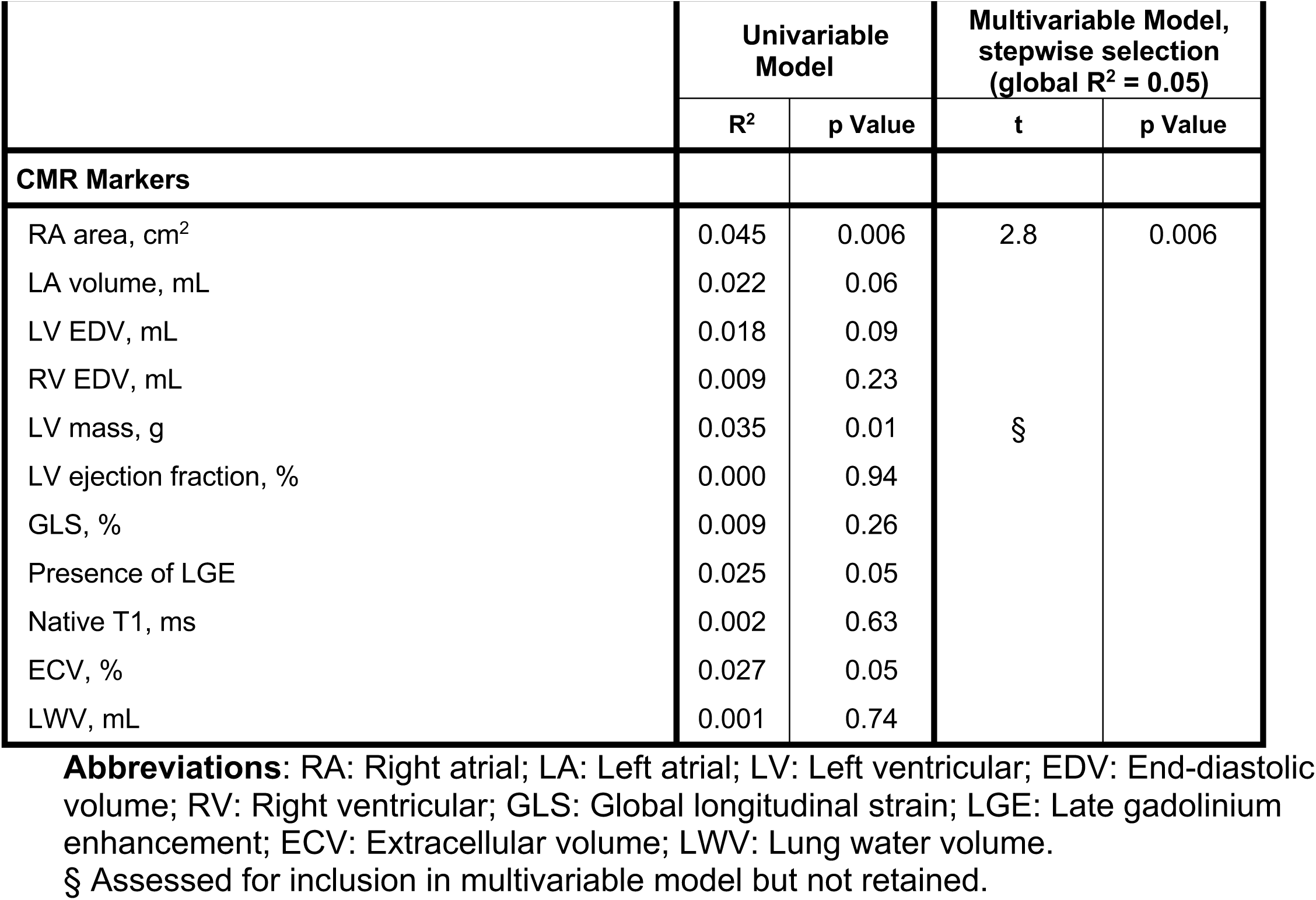
Univariable and multivariable linear regression models of CMR variables associated with LWD. Stepwise selection identified variables associated with LWD with p value <0.05 to enter and remain in the model.

|  | Univariable Model |  | Multivariable Model, stepwise selection (global R<sup>2</sup> = 0.05) |  |
| --- | --- | --- | --- | --- |
|  | R<sup>2</sup> | p Value | t | p Value |
| **CMR Markers** |  |  |  |  |
| RA area, cm<sup>2</sup> | 0.045 | 0.006 | 2.8 | 0.006 |
| LA volume, mL | 0.022 | 0.06 |  |  |
| LV EDV, mL | 0.018 | 0.09 |  |  |
| RV EDV, mL | 0.009 | 0.23 |  |  |
| LV mass, g | 0.035 | 0.01 | § |  |
| LV ejection fraction, % | 0.000 | 0.94 |  |  |
| GLS, % | 0.009 | 0.26 |  |  |
| Presence of LGE | 0.025 | 0.05 |  |  |
| Native T1, ms | 0.002 | 0.63 |  |  |
| ECV, % | 0.027 | 0.05 |  |  |
| LWV, mL | 0.001 | 0.74 |  |  |

**Abbreviations:** RA: Right atrial; LA: Left atrial; LV: Left ventricular; EDV: End-diastolic volume; RV: Right ventricular; GLS: Global longitudinal strain; LGE: Late gadolinium enhancement; ECV: Extracellular volume; LWV: Lung water volume.
§ Assessed for inclusion in multivariable model but not retained.

## Variables associated with LWV

Univariable and multivariable analyses for LWV are shown in Table 4. In univariable regression analysis, LWV was associated with age, male sex, BSA, LA volume, LV mass, LV end diastolic volume, RA area, right ventricular (RV) end diastolic volume and GLS (Figure 6). In a stepwise multivariable linear regression analysis including clinical and CMR variables, male sex, LA volume and RV end diastolic volume remained associated with LWV (global R^2^ = 0.40, p<0.001).

**Figure 6.**
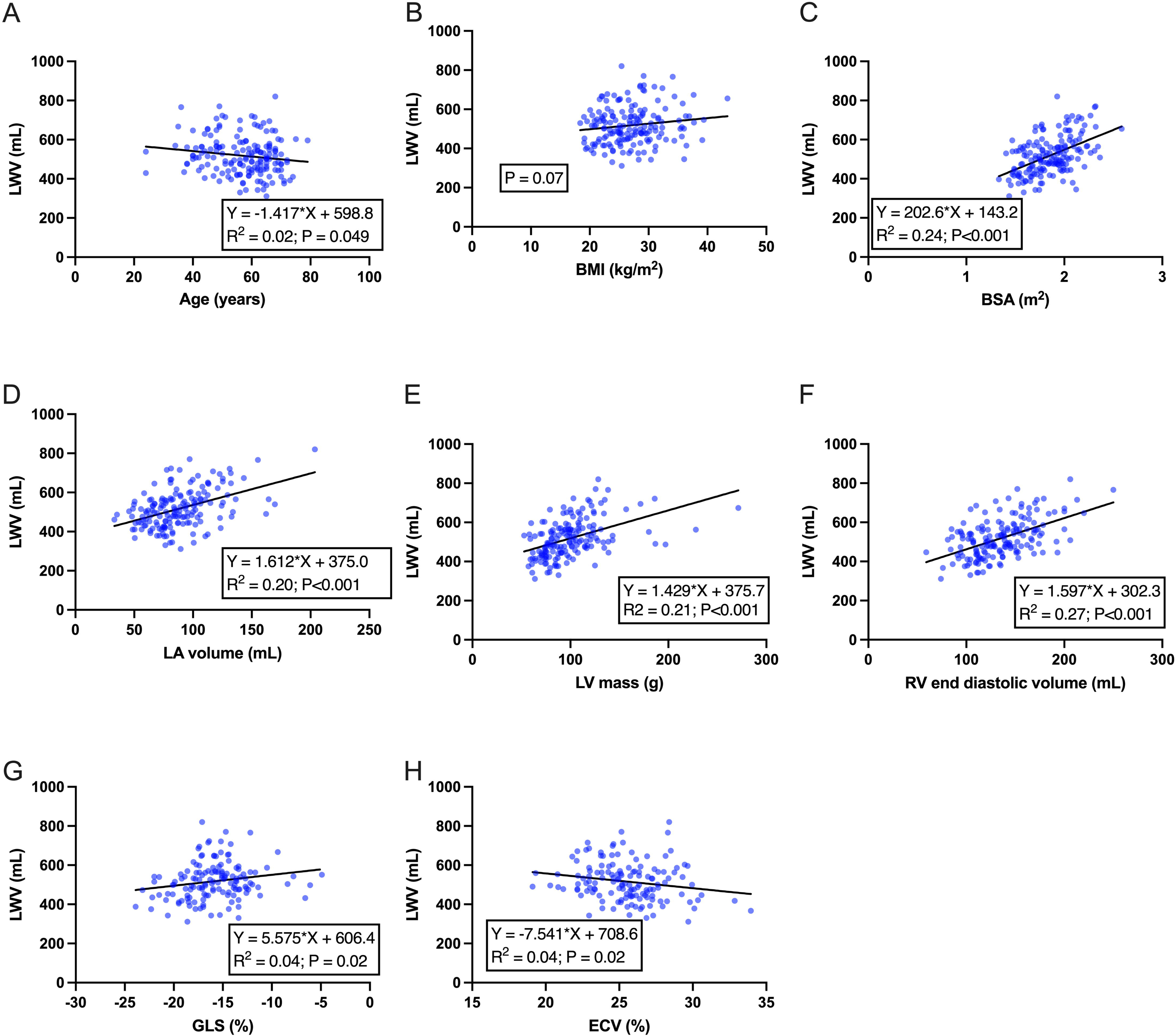
Scatter plots showing univariable relationships between lung water volume and clinical or routinely acquired CMR markers. Lung water volume was significantly associated with age (Panel A), BSA (Panel C), LA volume (Panel D), LV mass (Panel E), RV end diastolic volume (Panel F), GLS (Panel G) and ECV (Panel H). The line of best fit is shown for each analysis. Abbreviations: LWV: Lung water volume; BMI: Body mass index; BSA: Body surface area; LA: Left atrial; LV: Left ventricular; RV: Right ventricular; GLS: Global longitudinal strain; ECV: Extracellular volume fraction.

**Table 4.**
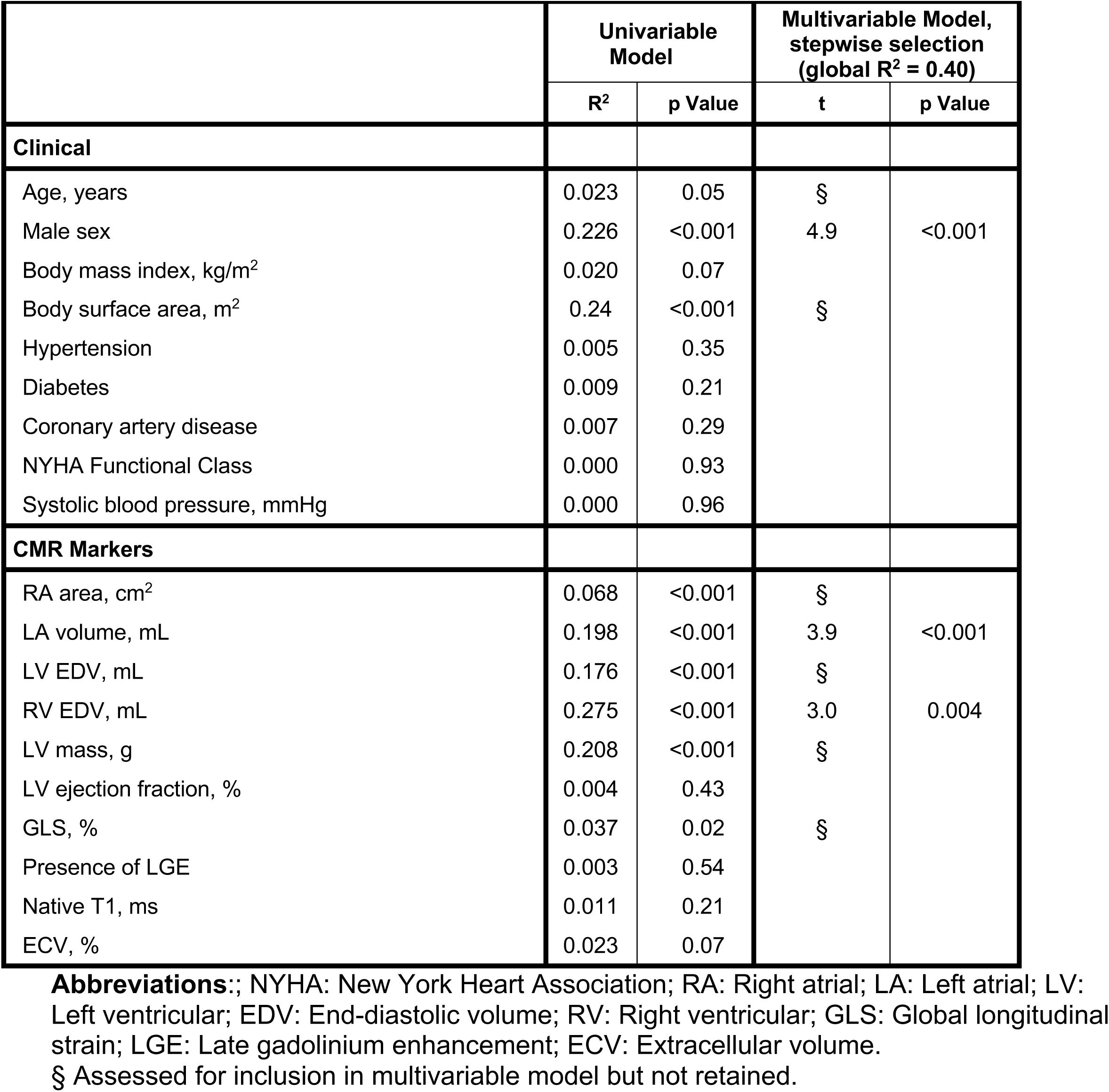
Univariable and multivariable linear regression models of variables associated with LWV. Stepwise selection identified variables associated with LWV with p value <0.05 to enter and remain in the model.

## Discussion

The main finding of this study is that LWD by CMR provides reasonable diagnostic performance for identifying patients at risk for heart failure, and is poorly correlated with all other routinely acquired CMR markers of disease. Of the CMR markers of cardiac size, mass, function and fibrosis tested, only RA area and LV mass were weakly correlated with LWD in univariable analyses. CMR-derived LWD has previously been shown to be strongly correlated with invasively-measured left-sided filling pressures and B-type natriuretic peptide [5].

Although pulmonary capillary wedge pressure can also be estimated using routine CMR markers using left atrial volume and left ventricular mass, correlation with invasively derived filling pressures at right heart catheterisation appear to be moderate at best [13, 14]. Therefore, CMR-derived LWD appears to provide important information regarding pulmonary congestion, possibly related to left-sided ventricular filling pressures, beyond what can currently be determined with conventional CMR markers.

The weak relationship observed between LWD and conventional CMR markers can be explained by the underlying pathophysiology of pulmonary oedema. Pulmonary oedema occurs in heart failure when LA pressure exceeds a critical threshold, resulting in increased pulmonary capillary hydrostatic pressure [1]. That threshold is variable, depending on the presence of a number of pulmonary adaptations, including reduced capillary filtration due to pulmonary vascular remodelling and endothelial dysfunction, enhanced alveolar fluid clearance and increased lymphatic drainage [15–18]. In some individuals, pulmonary capillary stress failure may occur due to pressure injury of the capillary wall and in the setting of pulmonary inflammation, resulting in alveolar flooding and oedema [19]. Therefore, the relationship between LA pressure elevation and pulmonary congestion is not straightforward and likely substantially modified by the physiological regulation of the pulmonary vascular system, which is not well accounted for in conventional CMR. CMR-derived LWD therefore appears to provide complementary information to routinely acquired CMR markers. Further work is required to determine whether LWD-guided management strategies can improve cardiovascular outcomes in heart failure beyond current algorithms.

The 3D Cartesian gradient-echo sequence used in this study to assess LWD has several advantages over the previously reported HASTE technique [2, 5, 20, 21]. The HASTE acquisitions use individual two-dimensional (2D) slices, which provide incomplete coverage of the lungs. HASTE is a spin-echo sequence with inherent T2-weighing and thus potentially increased or decreased signal intensities without a change in water density. The HASTE sequence uses a highly asymmetric k-space (half-Fourier technique) which can result in image artifacts and cause spatial variations in signal that can confound water quantification. The use of a single user-selected liver region as reference tissue to convert MRI signals to water density with the HASTE approach will be prone to systematic errors due to coil shading effects that cause spatial heterogeneity in the MRI signals, independent of water density.

These limitations are largely overcome by the sequence used in this study. 3D full torso imaging provides complete coverage of the lungs and all surrounding solid tissues. The reference signal used is a composite of all background signals, including the liver, heart and skeletal muscle, is user-independent and includes a signal normalization to correct for coil shading effects. A low flip angle (1 degree) eliminates T1 weighting [2]. The automated segmentation enables removal of confounding tissues and conduit blood vessels. Furthermore, a built-in respiratory navigator enables automated end-expiration reconstruction with free-breathing acquisitions.

The LWD values obtained in this study were higher compared to previously reported lung water density values using the HASTE sequence [5, 20]. This difference relates primarily to the differing methods used for background correction and normalization. In the previous studies, the liver was used as reference tissue and was assumed to have a water density of 70%. A similar correction was not applied in this study, in which the absolute water density in all of the surrounding tissues is acknowledged as unknown and assigned a reference value of 100%, and thus the lung water density values are relative to the composite multi-tissue reference. Other reasons for systematic differences in LWD between the two techniques could also include differing amounts of T2, T2* and T1 weighting [2, 5]. Regardless of these systematic differences, LWD continued to demonstrate acceptable discrimination between healthy controls and patients at risk of heart failure in our cohort, using a cut-off value of 27.6%, suggesting diagnostic utility.

In this study, LWD was most strongly associated with BMI. It is increasingly recognised that obesity is often associated with increased circulating blood volume and cardiac output, pulmonary venous and subsequently arterial hypertension, and ultimately, with left ventricular dysfunction and pulmonary oedema [22, 23]. Therefore, the relationship observed is in keeping with our current understanding of the pathophysiology of obesity. LWD was associated with LV mass in univariable regression analysis and the association with LA volume trended towards significance. This is in keeping with a recent CMR study which showed that of the routinely measured CMR metrics, only LV mass and LA volume were independently associated with invasively measured pulmonary capillary wedge pressure [13]. Given that LWD is also known to be associated with invasively derived filling pressures, this result is expected. Of the CMR markers assessed in this study, RA area showed the strongest association with LWD. RA area at echocardiography has been shown to distinguish healthy subjects from patients with RV volume overload, and may therefore serve as an early marker of pulmonary hypertension or other right-sided heart disease [24–26]. The association between LWD and RA area raises the possibility that LWD may be a sensitive, non-invasive marker for pulmonary hypertension. This could be investigated in future with CMR LWD and concurrent right heart catheterisation in a dedicated cohort with established pulmonary hypertension.

Lung water volume (LWV) showed the strongest association with markers of patient size, including sex and BSA. This is in keeping with the fact that extravascular lung water measured using the transpulmonary thermodilution technique is known to be size-dependent and is indexed to either actual or predicted body weight in clinical use [27, 28]. LA volume was independently associated with LWV in multivariable regression analysis, suggesting that LWV, in addition to LWD, may be associated with left ventricular filling pressures. RV end diastolic volume is known to be a marker of cardiac preload [29], which in turn is affected by volume status. The association between RV end diastolic volume and LWV may therefore mechanistically result from increased fluid accumulation in the lungs in the setting of increased pulmonary vascular hydrostatic pressure due to an increased circulating blood volume. LWV was not able to discriminate between the healthy control group and patients at risk of heart failure in this cohort. This was likely due, in part, to the dependence of this measure on the size of the lungs and/or the whole body. Follow-up work should assess the utility of CMR-derived non-invasive LWV indexed to body size in cohorts with and without heart failure to determine whether this measure has clinical utility.

### Limitations

In this study, we were not able to correlate the LWD results with B-type natriuretic peptide levels, a non-invasive marker of left ventricular filling pressures, or invasively measured pulmonary capillary wedge pressure. The cohort assessed in this study included patients undergoing CMR for clinical indications or as part of other research studies at our institution and these additional tests were therefore not included. Our cohort was overall relatively well and therefore only 2 patients met a combined outcome of hospitalisation for heart failure or death over a median follow-up time of 20 months, limiting the ability to perform any meaningful survival analysis. The 3D Cartesian pulse sequence had a moderately short TE of 0.43 ms, and thus still some T2* weighting in the lung parenchyma signal intensities.

## Conclusion

Lung water density, as measured using an updated 3D Cartesian gradient echo sequence at 1.5T, overcomes a number of the limitations of the older HASTE sequences and is able to discriminate between healthy controls and patients at risk of heart failure. Lung water density is only weakly explained by routinely acquired CMR parameters.

## Authors’ contributions

NRI, CWLC and MU designed the study. NRI, JAB and TTL collected the data. NRI and JGG analysed the data. NRI, CWLC and MU interpreted the data and wrote the manuscript. RBT made substantial revisions to the manuscript. All authors read and approved the final manuscript.

## Funding

None.

## Availability of data and materials

The datasets generated and analysed for the current study are not publicly available. Please contact the corresponding author for data requests.

## Ethics approval and consent to participate

Ethics approval was obtained from the Singhealth Centralized Institutional Review Board in Singapore, and all participants provided written informed consent. The study was conducted in accordance with the principles of the Declaration of Helsinki.

## Disclosures

MU is principal investigator for an institutional research and development agreement regarding cardiovascular magnetic resonance imaging between Karolinska University Hospital and Siemens. All other authors have reported that they have no relationships relevant to the contents of this paper to disclose.

## Notes

### Funding Statement

This study did not receive any funding

### Author Declarations

Ethics committee/IRB of Singapore Health Services (SingHealth) gave ethical approval for this work

